# Baseline and Serial BNP Measurements Predict Long-Term Mortality in Adults with Congenital Heart Disease: A study including 3,392 patients

**DOI:** 10.1101/2023.08.24.23294392

**Authors:** Yusuke Yumita, Zhuoyuan Xu, Gerhard-Paul Diller, Aleksander Kempny, Isma Rafiq, Claudia Montanaro, Wei Li, Koichiro Niwa, Hong Gu, Konstantinos Dimopoulos, Michael A Gatzoulis, Margarita Brida

## Abstract

**Background:** Although there is an ever growing number of adult patients with congenital heart disease (ACHD), many are still afflicted by premature death. Previous reports suggested that natriuretic peptides may identify ACHD patients with adverse outcome. We investigated the prognostic power of brain natriuretic peptide (BNP) across the spectrum of ACHD in a large contemporary cohort.

**Methods:** We retrospectively studied 3,392 consecutive and well-characterised ACHD patients under long-term follow-up at a tertiary ACHD centre between 2006-2019. The primary study endpoint was all-cause mortality.

**Results:** A total of 11,974 BNP measurements were analysed. The median BNP at baseline was 47 [24-107] ng/L. During a median follow-up of 8.6 years (29,115 patient-years), 615 (18.1%) patients died. On univariate and multivariate analysis both baseline BNP and temporal changes in BNP levels were predictive of mortality (p<0.001 for both) independent of congenital heart disease diagnosis, complexity, anatomic/haemodynamic features, and/or systolic systemic ventricular function. Patients within the highest quartile of baseline BNP (>107 ng/L) and those within the highest quartile of temporal BNP change (>35 ng/L) had 5.8 and 3.6-fold increased risk of death, respectively.

**Conclusion:** Baseline BNP and temporal BNP changes are both significantly associated with all-cause mortality in ACHD independent of congenital heart disease diagnosis, complexity, anatomic/haemodynamic features, and/or systolic systemic ventricular function. BNP levels represent an easy to obtain and inexpensive marker conveying prognostic information and should be used for the routine surveillance of patients with ACHD.

## Introduction

Continuous improvements in the diagnosis and treatment of congenital heart disease (CHD) have brought about an ever-growing population of adults with CHD (ACHD), increasingly including older patients, all with unique healthcare needs. However, ACHD patients are still afflicted by premature death, with heart failure being the leading cause of morbidity and mortality.^1^ This has important implications, including increasing rates of hospitalisation, higher healthcare utilisation, and higher overall health resource use and costs.^2, 3^ Risk stratification of all patients at the time of diagnosis and during periodic life-long follow-up is a fundamental part of ACHD practice, aimed at identifying patients at increased risk of adverse outcomes, who require escalation of treatment and closer surveillance.

Natriuretic peptides, either brain natriuretic peptide (BNP) or N-terminal pro-B-type natriuretic peptide (NTproBNP), are established biomarkers in acquired heart failure, used both for diagnosis and assessment of disease severity course. Natriuretic peptide levels and their change over time are also important predictors of heart failure hospitalization and mortality.^4, 5^

Similar to acquired heart disease, natriuretic peptides are considered clinically relevant in ACHD.^6, 7^ Previous studies have shown that an elevated single natriuretic peptide value^6^ and its increase over time is associated with the risk of adverse events in ACHD.^7-10^ However, variability in their levels across the spectrum of different CHD with different underlying cardiac abnormalities and haemodynamics, exercise limitation, and degree of neurohormonal activation^9, 11^ limit their implementation as a screening tool for such a heterogenous population. The aim of our study was, therefore, to examine the prognostic role of BNP and its serial measurements in a large, contemporary, and well-characterized cohort of ACHD patients.

## Methods

We retrospectively examined data on 3,392 consecutive, clinically stable ACHD patients under periodic long-term follow-up at our large tertiary ACHD centre (Royal Brompton Hospital, London) who had BNP measurements performed between 2006 and 2019. We studied patients’ data including demographics, primary diagnosis, previous interventions, clinical characteristics, laboratory parameters, echocardiographic findings, and cardiopulmonary exercise testing from dedicated electronic health records.^12^ Exclusion criteria were defined as age < 18 years, acute decompensated heart failure, current systemic infection, or malignancy at the time of BNP sampling, severe renal impairment (serum creatinine level >200 umol/l) and/or liver dysfunction (liver aspartate transaminase more than twice the upper limit of normal (>82 IU/L). Laboratory, echocardiographic, and cardiopulmonary exercise data were linked to clinical visits within 6 months of the respective tests. Systemic ventricular function was assessed by echocardiography and graded in a semi-quantitative manner into normal, mildly, moderately, or severely impaired. Patients were manually categorised according to I) CHD diagnosis, II) disease complexity based on the established Bethesda classification, III) distinct morphologic and hemodynamic features (univentricular heart, systemic right ventricle, Eisenmenger syndrome, Tetralogy of Fallot spectrum, and remaining others with systemic left ventricle who were defined as the reference group). ^9, 13 14^ Cyanosis was defined as oxygen saturations <90% at rest. New York Heart Association (NYHA) functional class and medications were manually extracted from the clinical records. Heart failure medications were defined by combination usage of more than one of the following medications: β-blocker, angiotensin converting enzyme inhibitor (ACEI) or angiotensin receptor blocker (ARB) or angiotensin receptor neprilysin inhibitor (ARNI), mineralocorticoid antagonist (MRA), and diuretics. The temporal change of BNP levels (ΔBNP) was defined as the difference of BNP level between one measurement level minus the previous BNP level. Data on overall mortality were retrieved from the Office for National Statistics, which registers all United Kingdom deaths, and all-cause mortality served as the study primary endpoint. As this was a retrospective analysis based on data collected for routine clinical care and administrative purposes (UK National Research Ethics Service guidance), individual informed consent was not required. The study was approved by the Institutional Research and Governance Committee.

### Statistical Analysis

Continuous variables are presented as mean ± standard deviation (SD) when normally distributed or as median with interquartile range (IQR) if the distribution departed from normality, whereas categorical variables are presented as numbers (%). Comparison of continuous variables between groups was performed using the non-parametric Wilcoxon rank sum test or Kruscal–Wallis test, while categorical variables were compared using the Fisher’s exact test. For the prognostic model, overall survival data was included in univariate and multivariate time dependent Cox proportional hazard survival models after testing the proportional hazards assumption (by assessing the relationship between scaled Schoenfeld residuals and survival time). To assess the effect of change in BNP after the first BNP measurement, stratified by outcome, a random effects mixed linear model was employed using the lme4 and lmerTest packages. Analyses were performed using R-package version 4.1.0. A two-sided P-value of <0.05 was considered indicative of statistical significance.

## Results

### I. Study population

Baseline demographics, clinical characteristics, and laboratory data are summarised in **Table 1**; during the study period, 3,392 ACHD patients (male n=1714, 50.5%) had at least one BNP measurement. The mean age at first BNP measurement (baseline) was 40.0 ± 17.0 years, whereas median BMI was 24.5 kg/m^2^ (IQR 21.9-28.3). Moreover, 2,509 (74.0%), 665 (19.6%), and 218 (6.4%) patients were in the NYHA functional class D, D, and D/D, respectively. There were 1,450 (42.7%) patients with simple, 1,225 (36.1%) moderate, and 717 (21.1%) with complex CHD. During a median follow-up time of 8.6 years (IQR 5.9-11.0, corresponding to a total of 29,115 patient-years of follow-up), 615 (18.1%) patients died.

**Table 1.** The baseline characteristics of the study population.

| Main diagnosis | Number | Age (years)<br>Median [IQR] | Male (%) | BMI (kg/m<sup>2</sup>) | SBP (mmHg) | Sinus rhythm (%) | Cyanosis (%) | NYHA class □, □, and □/□ (%) | Creatinine (umol/L) | BNP (pg/mL)<br>Median [IQR] | Systemic ventricular function<br>normal/mild/moderate/severe (%) | Deceased (%) |
| --- | --- | --- | --- | --- | --- | --- | --- | --- | --- | --- | --- | --- |
| APVC | 21 | 31 [25-57] | 38% | 26.0±5.8 | 117±14 | 95% | 0.0% | 86/14/0 | 71.0±18.9 | 32 [21-76] | 89/11/0/0 | 24% |
| ASD/PFO | 368 | 47 [32-60] | 45% | 26.7±5.8 | 123±19 | 85% | 2.1% | 74/22/4 | 76.5±20.9 | 42 [18-121] | 87/8/4/1 | 21% |
| AVSD | 109 | 36 [27-48] | 38% | 26.4±5.9 | 117±18 | 82% | 0.9% | 85/12/3 | 72.9±15.3 | 57 [33-116] | 86/8/4/2 | 10% |
| CCTGA | 73 | 41 [29-54] | 55% | 24.7±4.7 | 114±15 | 84% | 8.2% | 64/30/5 | 79.1±17.4 | 103 [30-272] | 41/14/31/14 | 19% |
| CoA | 328 | 36 [25-47] | 55% | 25.8±5.2 | 127±17 | 95% | 0.6% | 90/9/1 | 75.7±18.4 | 28 [15-64] | 90/6/2/2 | 6% |
| DORV | 50 | 31 [22-38] | 56% | 24.5±5.6 | 115±12 | 88% | 8.0% | 74/20/6 | 74.2±18.7 | 40 [20-105] | 73/10/17/0 | 18% |
| Ebstein | 110 | 41 [27-52] | 39% | 25.9±5.7 | 118±17 | 94% | 1.8% | 85/11/4 | 73.1±19.4 | 44 [24-102] | 89/8/0/3 | 5% |
| Eisenmenger | 243 | 35 [27-44] | 32% | 25.2±6.4 | 116±18 | 96% | 68% | 12/46/43 | 81.8±22.4 | 55 [26-128] | 87/9/3/1 | 49% |
| Fontan | 48 | 28 [22-35] | 54% | 23.4±4.4 | 114±13 | 94% | 21% | 60/38/2 | 76.4±21.5 | 45 [23-120] | 70/21/5/4 | 17% |
| Heterotaxy | 19 | 33 [26-39] | 32% | 20.8±3.9 | 110±13 | - | 53% | 63/26/11 | 67.7±13.2 | 69 [50-107] | 79/21/0/0 | 26% |
| Marfan | 55 | 41 [32-55] | 53% | 24.5±4.9 | 119±17 | 98% | 1.8% | 91/9/0 | 71.6±18.9 | 60 [31-156] | 76/17/5/2 | 18% |
| Pulmonary atresia | 60 | 28 [23-43] | 47% | 23.4±5.2 | 112±17 | 98% | 38% | 57/28/15 | 74.3±23.7 | 62 [31-130] | 71/21/4/4 | 28% |
| PDA | 27 | 43 [24-61] | 30% | 25.9±7.6 | 117±19 | 89% | 3.7% | 74/26 | 69.6±15.9 | 55 [27-194] | 73/9/14/4 | 11% |
| TOF | 521 | 32 [24-45] | 53% | 25.4±5.5 | 117±15 | 93% | 1.7% | 86/12/2 | 76.1±19.2 | 42 [24-78] | 87/9/3/1 | 9% |
| TGA arterial switch or Rastelli | 70 | 22 [19-28] | 60% | 24.8±5.3 | 119±17 | 94% | 0.0% | 93/7/0 | 75.0±15.4 | 34 [21-46] | 81/18/1/0 | 1% |
| TGA atrial switch | 103 | 32 [29-40] | 59% | 25.1±4.5 | 114±13 | 87% | 5.8% | 70/25/5 | 75.0±17.6 | 62 [35-114] | 81/10/6/3 | 17% |
| Truncus arteriosus | 18 | 26 [21-35] | 61% | 22.7±4.2 | 120±16 | 100% | 17% | 83/11/6 | 70.7±14.6 | 45 [17-119] | 87/13/0/0 | 17% |
| Univentricular heart | 86 | 30 [24-41] | 56% | 23.3±5.0 | 114±15 | 78% | 48% | 40/38/22 | 81.3±26.1 | 91 [39-161] | 57/26/14/3 | 26% |
| Valvar disease | 823 | 45 [30-61] | 55% | 26.3±5.3 | 124±18 | 89% | 1.9% | 77/20/3 | 81.2±25.3 | 50 [24-125] | 88/9/2/1 | 24% |
| VSD | 232 | 33 [24-45] | 56% | 25.7±5.2 | 122±18 | 94% | 0.8% | 82/16/2 | 74.8±21.0 | 37 [17-86] | 83/13/2/2 | 10% |
| Other | 28 | 39 [27-60] | 39% | 25.8±5.9 | 119±13 | 93% | 0.0% | 89/7/4 | 72.5±12.3 | 61 [26-106] | 63/21/16/0 | 11% |
| Total | 3392 | 37 [26-50] | 51% | 25.7±5.5 | 120±18 | 91% | 9% | 74/20/6/0 | 77.3±21.4 | 47 [24-107] | 84/10/4/2 | 18% |

APVC, anomalous pulmonary venous connection; ASD, atrial septal defect; PFO, patent foramen ovale; AVSD, atrioventricular septal defect; BMI, body mass index; BNP, B-type natriuretic peptide; CCTGA, congenitally corrected transposition of the great arteries; CoA spectrum, coarctation of the aorta or interrupted aortic arch; DORV, double outlet right ventricle; NYHA, New York Heart Association functional class; PDA, patent ductus arteriosus; SBP, systolic blood pressure; TOF, Tetralogy of Fallot; TGA, transposition of the great arteries, VSD, ventricular septal defect. Other includes anomalous coronary artery, sinus of Valsalva aneurysm and double chambered right ventricle

### II. BNP and ΔBNP

Overall, a total of 11,974 BNP measurements were available in the study population with a median BNP of 47 ng/L (IQR 24-107 ng/L). Amongst the 3,392 overall patients with BNP assessment, 2,020 (59.6%) patients had multiple BNP measurements during the study period. The median ΔBNP was 3.5 ng/L (IQR -20 to 35 ng/L) and the median time interval between BNP measurements was 189 days (IQR 89-460 days). The distribution of BNP at baseline and the distribution of ΔBNP is shown in **Supplementary** Figure 1. There was a significant correlation between BNP levels and NYHA functional class, as well as systemic ventricular function impairment (both p<0.001). Higher BNP levels were seen with increasing disease severity, including worse NYHA functional class and more impaired systemic ventricular function, as shown in **Figure 1A and 1B**. In contrast, ΔBNP levels did not significantly correlate with change in functional class (p=0.74), but did correlate with worsening systemic ventricular function (p=0.002) as shown in **Figure 1C and 1D**.

### III. Prognostic value of BNP and ΔBNP

*BNP at baseline*. On univariate Cox analysis age (HR 1.05, 95% CI 1.05-1.06), higher NYHA functional class (NYHA D-D; HR 3.58, 95% CI 3.05-4.21), cyanosis (HR 2.76, 95% CI 2.29-3.33), creatinine concentration (HR 1.02, 95% CI 1.02-1.03), systemic ventricular dysfunction (more than mildly reduced systolic function; HR 2.95, 95% CI 2.19-3.96), treatment with a β-blocker (HR 1.63, 95% CI 1.45-1.83), or heart failure medication (HR 3.59, 95% CI 3.06-4.20), and BNP (HR per /100-ng/L increase 1.16, 95% CI 1.15-1.18) were significantly associated with a higher risk of all-cause mortality (p<0.001 for all). Disease complexity (HR 1.67, 95% CI 1.41-1.97, p=0.001) and Eisenmenger syndrome (HR 2.02, 95% CI 1.70-2.61, p<0.001) were also associated with a higher mortality risk (**Table 2**).

**Table 2.**
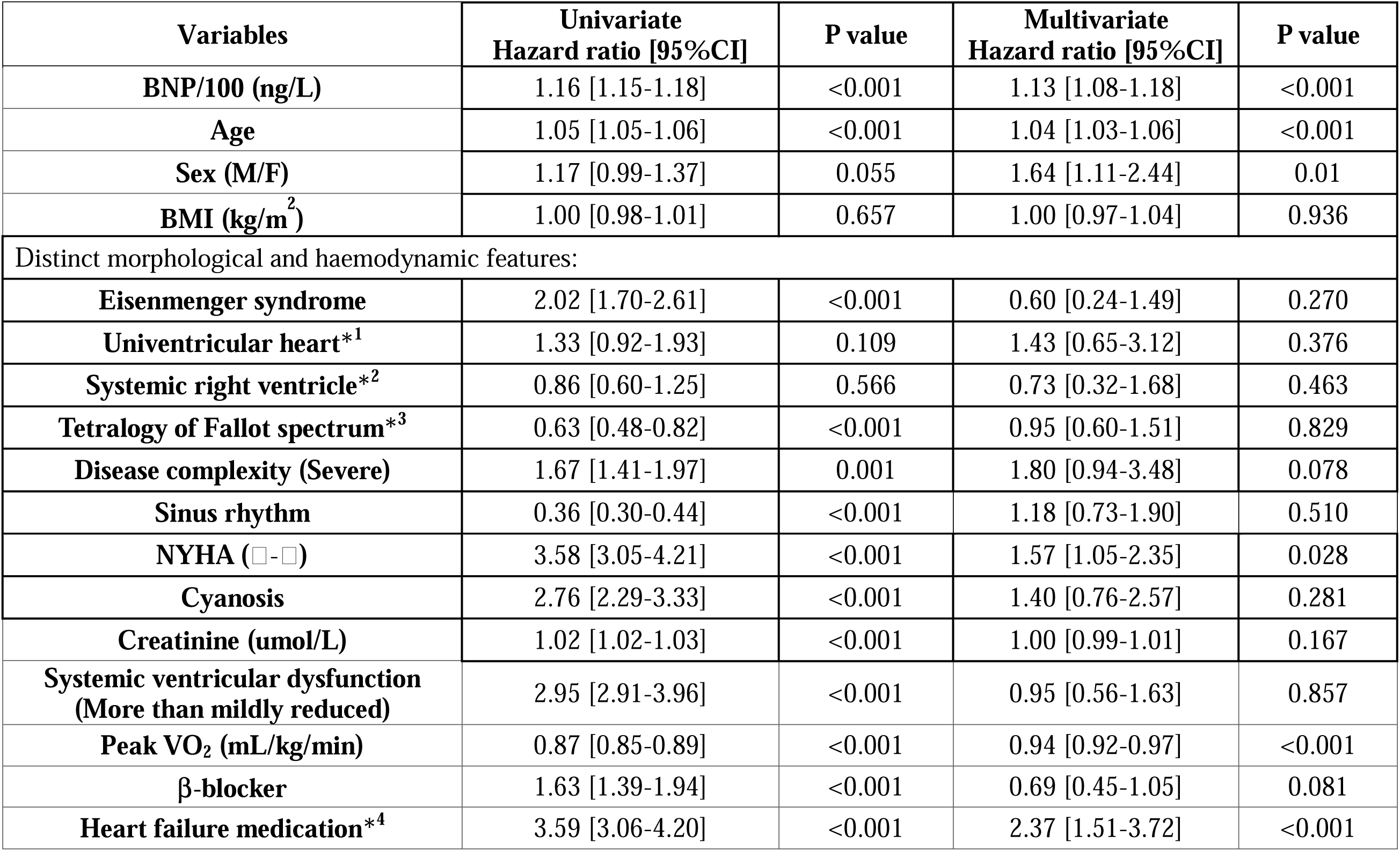
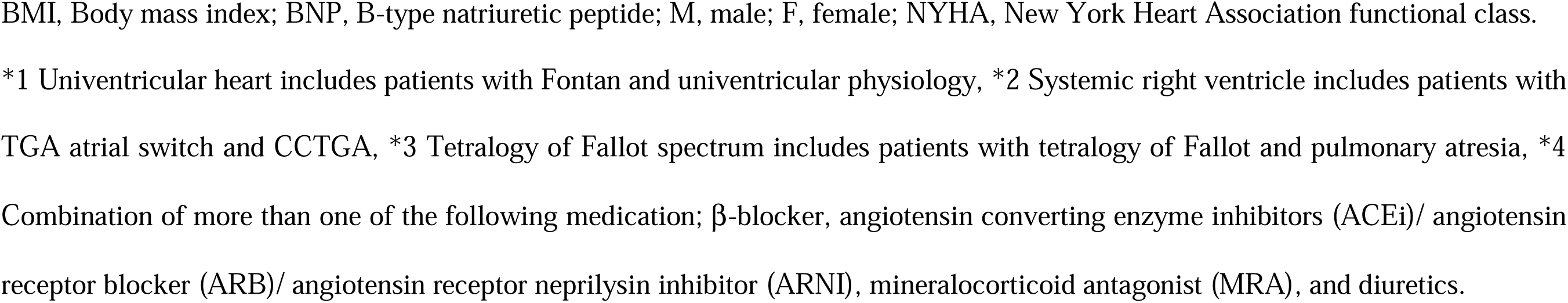
Cox hazard analysis based on baseline BNP.

| **Variables** | **Univariate**<br>**Hazard ratio [95%CI]** | **P value** | **Multivariate**<br>**Hazard ratio [95%CI]** | **P value** |
| --- | --- | --- | --- | --- |
| **BNP/100 (ng/L)** | 1.16 [1.15-1.18] | <0.001 | 1.13 [1.08-1.18] | <0.001 |
| **Age** | 1.05 [1.05-1.06] | <0.001 | 1.04 [1.03-1.06] | <0.001 |
| **Sex (M/F)** | 1.17 [0.99-1.37] | 0.055 | 1.64 [1.11-2.44] | 0.01 |
| **BMI (kg/m<sup>2</sup>)** | 1.00 [0.98-1.01] | 0.657 | 1.00 [0.97-1.04] | 0.936 |
| **Distinct morphological and haemodynamic features:** |  |  |  |  |
| **Eisenmenger syndrome** | 2.02 [1.70-2.61] | <0.001 | 0.60 [0.24-1.49] | 0.270 |
| **Univentricular heart\*<sup>1</sup>** | 1.33 [0.92-1.93] | 0.109 | 1.43 [0.65-3.12] | 0.376 |
| **Systemic right ventricle\*<sup>2</sup>** | 0.86 [0.60-1.25] | 0.566 | 0.73 [0.32-1.68] | 0.463 |
| **Tetralogy of Fallot spectrum\*<sup>3</sup>** | 0.63 [0.48-0.82] | <0.001 | 0.95 [0.60-1.51] | 0.829 |
| **Disease complexity (Severe)** | 1.67 [1.41-1.97] | 0.001 | 1.80 [0.94-3.48] | 0.078 |
| **Sinus rhythm** | 0.36 [0.30-0.44] | <0.001 | 1.18 [0.73-1.90] | 0.510 |
| **NYHA (□-□)** | 3.58 [3.05-4.21] | <0.001 | 1.57 [1.05-2.35] | 0.028 |
| **Cyanosis** | 2.76 [2.29-3.33] | <0.001 | 1.40 [0.76-2.57] | 0.281 |
| **Creatinine (umol/L)** | 1.02 [1.02-1.03] | <0.001 | 1.00 [0.99-1.01] | 0.167 |
| **Systemic ventricular dysfunction**<br>**(More than mildly reduced)** | 2.95 [2.91-3.96] | <0.001 | 0.95 [0.56-1.63] | 0.857 |
| **Peak VO<sub>2</sub> (mL/kg/min)** | 0.87 [0.85-0.89] | <0.001 | 0.94 [0.92-0.97] | <0.001 |
| **β-blocker** | 1.63 [1.39-1.94] | <0.001 | 0.69 [0.45-1.05] | 0.081 |
| **Heart failure medication\*<sup>4</sup>** | 3.59 [3.06-4.20] | <0.001 | 2.37 [1.51-3.72] | <0.001 |

BMI, Body mass index; BNP, B-type natriuretic peptide; M, male; F, female; NYHA, New York Heart Association functional class.
\*1 Univentricular heart includes patients with Fontan and univentricular physiology, \*2 Systemic right ventricle includes patients with TGA atrial switch and CCTGA, \*3 Tetralogy of Fallot spectrum includes patients with tetralogy of Fallot and pulmonary atresia, \*4 Combination of more than one of the following medication; β-blocker, angiotensin converting enzyme inhibitors (ACEi)/ angiotensin receptor blocker (ARB)/ angiotensin receptor neprilysin inhibitor (ARNI), mineralocorticoid antagonist (MRA), and diuretics.

On multivariate Cox analysis, BNP independently predicted all-cause mortality after adjusting for confounding factors including age, distinct morphological and haemodynamic features, disease complexity, heart rhythm, NYHA functional class D-D, cyanosis, creatinine, systemic ventricular dysfunction, peak VO_2_, and prescription of β-blocker and heart failure medication (HR per 100-ng/L increase 1.13, 95% CI 1.08-1.18, p<0.001) (**Table 2**).

The estimated average baseline BNP at study inclusion (intercept) was significantly higher in patients who reached the primary endpoint during the study compared to the remainder (p<0.001), 274 (95%CI 255-293) ng/L vs 90 (95%CI 86-94) ng/L, respectively. Moreover, the average increase of BNP (ΔBNP slope) was significantly higher in patients who reached the primary endpoint compared to the remaining patients, 47 (95%CI 41-53) ng/L versus 3 (95%CI 2-4) ng/L (p<0.001) as presented in **Figure 2** based on the results of the mixed effects linear models. When stratifying patients by quartiles of BNP, the risk of all-cause mortality increased with higher BNP level as shown in **Figure 3A**. Patients within the highest quartile of BNP (>107 ng/L), had a hazard ratio of 5.77 (95% CI 4.91-6.79, p<0.001) of dying compared to the remaining patients (**Figure 4**).

*Serially assessed BNP.* On univariate time dependent Cox proportional hazard analysis age (HR 1.06, 95% CI 1.05-1.06), higher NYHA functional class (NYHA D-D; HR 3.47, 95% CI 2.80-4.31), disease complexity (HR 1.53, 95% CI 1.24-1.88), cyanosis (HR 2.68, 95% CI 2.15-3.35), creatinine concentration (HR 1.03, 95% CI 1.02-1.03), systemic ventricular dysfunction (more than mildly reduced systolic function; HR 2.99, 95% CI 2.18-4.10), prescription of heart failure medications (HR 3.37, 95% CI 2.74-4.14) and ΔBNP (HR per /Δ100-ng/L increase 1.22, 95% CI 1.19-1.26) were all significantly associated with a higher risk of all-cause mortality (p<0.001 for all). Eisenmenger syndrome (HR 1.84, 95% CI 1.43-2.37) and prescription of β-blocker (HR 1.27 95% CI 1.03-1.57) were also associated with a higher mortality risk (**Table 3**).

**Table 3.**
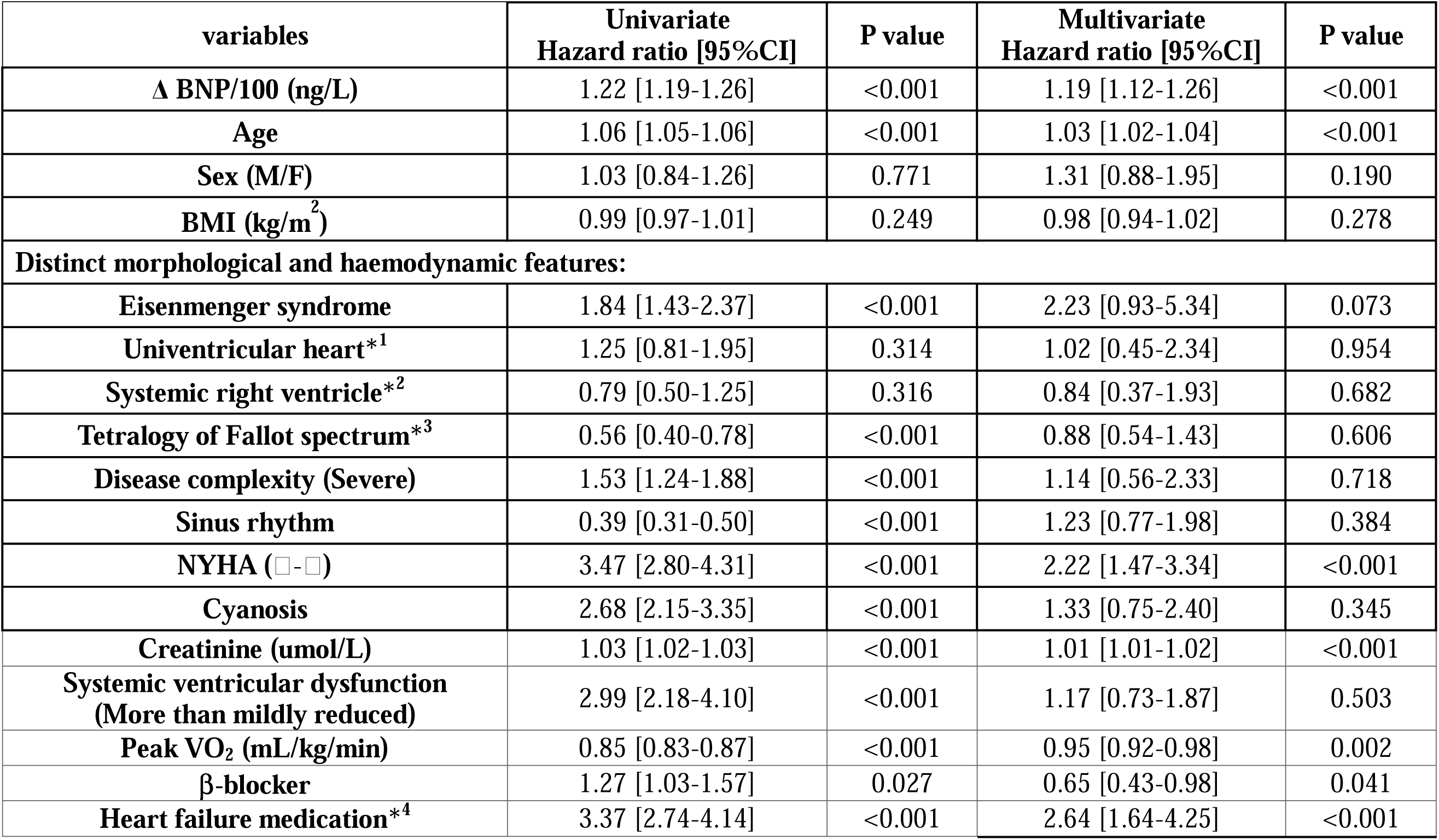
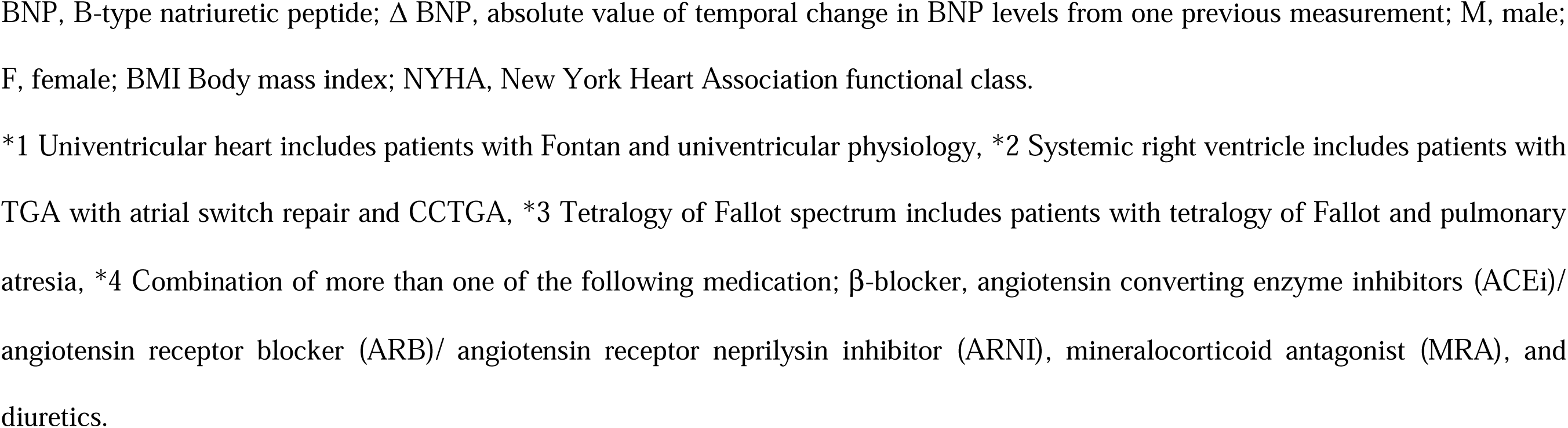
Time dependent cox proportional hazard analysis for serial BNP.

| **variables** | **Univariate**<br>**Hazard ratio [95%CI]** | **P value** | **Multivariate**<br>**Hazard ratio [95%CI]** | **P value** |
| --- | --- | --- | --- | --- |
| **Δ BNP/100 (ng/L)** | 1.22 [1.19-1.26] | <0.001 | 1.19 [1.12-1.26] | <0.001 |
| **Age** | 1.06 [1.05-1.06] | <0.001 | 1.03 [1.02-1.04] | <0.001 |
| **Sex (M/F)** | 1.03 [0.84-1.26] | 0.771 | 1.31 [0.88-1.95] | 0.190 |
| **BMI (kg/m<sup>2</sup>)** | 0.99 [0.97-1.01] | 0.249 | 0.98 [0.94-1.02] | 0.278 |
| **Distinct morphological and haemodynamic features:** |  |  |  |  |
| **Eisenmenger syndrome** | 1.84 [1.43-2.37] | <0.001 | 2.23 [0.93-5.34] | 0.073 |
| **Univentricular heart\*<sup>1</sup>** | 1.25 [0.81-1.95] | 0.314 | 1.02 [0.45-2.34] | 0.954 |
| **Systemic right ventricle\*<sup>2</sup>** | 0.79 [0.50-1.25] | 0.316 | 0.84 [0.37-1.93] | 0.682 |
| **Tetralogy of Fallot spectrum\*<sup>3</sup>** | 0.56 [0.40-0.78] | <0.001 | 0.88 [0.54-1.43] | 0.606 |
| **Disease complexity (Severe)** | 1.53 [1.24-1.88] | <0.001 | 1.14 [0.56-2.33] | 0.718 |
| **Sinus rhythm** | 0.39 [0.31-0.50] | <0.001 | 1.23 [0.77-1.98] | 0.384 |
| **NYHA (□-□)** | 3.47 [2.80-4.31] | <0.001 | 2.22 [1.47-3.34] | <0.001 |
| **Cyanosis** | 2.68 [2.15-3.35] | <0.001 | 1.33 [0.75-2.40] | 0.345 |
| **Creatinine (umol/L)** | 1.03 [1.02-1.03] | <0.001 | 1.01 [1.01-1.02] | <0.001 |
| **Systemic ventricular dysfunction**<br>**(More than mildly reduced)** | 2.99 [2.18-4.10] | <0.001 | 1.17 [0.73-1.87] | 0.503 |
| **Peak VO<sub>2</sub> (mL/kg/min)** | 0.85 [0.83-0.87] | <0.001 | 0.95 [0.92-0.98] | 0.002 |
| **β-blocker** | 1.27 [1.03-1.57] | 0.027 | 0.65 [0.43-0.98] | 0.041 |
| **Heart failure medication\*<sup>4</sup>** | 3.37 [2.74-4.14] | <0.001 | 2.64 [1.64-4.25] | <0.001 |

BNP, B-type natriuretic peptide; ΔBNP, absolute value of temporal change in BNP levels from one previous measurement; M, male; F, female; BMI Body mass index; NYHA, New York Heart Association functional class.
\*1 Univentricular heart includes patients with Fontan and univentricular physiology, \*2 Systemic right ventricle includes patients with TGA with atrial switch repair and CCTGA, \*3 Tetralogy of Fallot spectrum includes patients with tetralogy of Fallot and pulmonary atresia, \*4 Combination of more than one of the following medication; β-blocker, angiotensin converting enzyme inhibitors (ACEi)/ angiotensin receptor blocker (ARB)/ angiotensin receptor neprilysin inhibitor (ARNI), mineralocorticoid antagonist (MRA), and diuretics.

On multivariate time dependent Cox proportional hazard analysis ΔBNP independently predicted all-cause mortality (HR per Δ100ng/L increase 1.19, 95% CI 1.12-1.26, p<0.001) after adjusting for confounding factors including age, distinct morphological and haemodynamic features, disease complexity, heart rhythm, NYHA functional class D-D, cyanosis, creatinine, systemic ventricular dysfunction, peak VO_2_, and prescription of β-blocker and HF medication. Similar to BNP, patients within the highest quartile of ΔBNP (>35 ng/L) had the highest risk of mortality **(Figure 3B)** and a hazard ratio of 3.59 (95% CI 2.93-4.40, p<0.001) of death compared to the remaining (**Figure 5**).

## Discussion

We report herewith the largest study to date regarding baseline and serial BNP measurements in a contemporary cohort of 3,392 ACHD patients with 29,115 patient-years follow-up. Both isolated BNP measurements and temporal BNP change correlated closely with all-cause mortality independently of the underlying CHD diagnosis, disease complexity, specific anatomic features, and systolic ventricular function. Patients with a BNP value in the highest quartile of measurements had an approximately 6-fold increased mortality risk compared to the remainder. A similar effect was seen for change in BNP, where patients within the highest quartile of ΔBNP had 3.6-fold increased risk of death.

### BNP and underlying CHD

Our data supports earlier evidence linking natriuretic peptides levels with outcome in ACHD. ^7-11^ However, potential variability in BNP levels across the spectrum of different CHD diagnoses with different underlying cardiac abnormalities limited the implementation of these earlier reports into clinical practice.^13, 15^ In our, much larger ACHD contemporary cohort, we demonstrate unequivocally that both BNP and ΔBNP relate to all-cause mortality, independently of underlying congenital heart disease diagnoses, severity and morphological features in the very heterogeneous spectrum of ACHD. Furthermore, our data shows that serial BNP assessment is a valuable adjunct to absolute BNP measurements ^7^ and in combination, identifies the cohort at the highest risk of mortality.

### BNP and NYHA functional class

Patients with ACHD are afflicted by a life-long disease, with multiple adaptations occurring during their trajectory^16^. Nonetheless, the NYHA classification remains a valuable clinical tool with correlation to exercise capacity, severity of underlying cardiac disease and mid-to long-term mortality.^16^ Our data showed that baseline BNP values correlate well with NYHA functional class, thus identifying more symptomatic and higher-risk patients. Interestingly, however, there was no correlation with ΔBNP and change in NYHA class, albeit they well identify independently patients at risk of mortality. We submit, therefore, ΔBNP might be more sensitive than NYHA functional class in identifying patients at risk of clinical deterioration; ΔBNP might manifest itself before NYHA functional class change is appreciated, begging for additional medical attention and potential further therapeutic intervention/s.

### BNP and systemic ventricular function

ACHD patients have multiple factors influencing their prognosis and, for some, the progression of heart failure. In the follow-up assessment of these patients, we routinely employ standardized transthoracic echocardiography to assess ventricular function, an established prognostic marker.^17^ Our study demonstrated that baseline BNP levels and ΔBNP were significantly associated with systemic ventricular dysfunction. This is even more relevant in the complex and heterogenous underlying anatomy of CHD patients, as echocardiographic assessment of ventricular function is often not straightforward, requires tertiary expertise, and even then, subtle changes in function can be overlooked. Moreover, serial echocardiograms are time consuming, often subject to interobserver variability, and relatively costly. Cardiac magnetic resonance, the ‘gold standard’ for assessing systolic ventricular function in such a heterogeneous ACHD population is costly and not universally available, thus unlikely to serve as a routine follow up tool. Furthermore there is a real challenge in assessing diastolic function of the systemic, pulmonary, or single ventricle in such a heterogeneous group in a uniform way by any imaging modality, albeit this impacts on prognosis and outcome in ACHD cohorts.^18^ Hence the value of BNP and ΔBNP - as surrogate markers of global heart function - which our data suggest, are predictive of mortality and correlates with gross assessment of systemic ventricular function. We advocate, therefore, that periodic BNP measurements, being ‘objective’, inexpensive, and easy to obtain, should be employed in the serial assessment of patients at risk of heart failure and adverse outcome from within the ACHD cohort.

### Higher BNP and ΔBNP levels and increased mortality risk

Our study showed that patients with the highest BNP and the highest temporal change of BNP levels were at the highest risk of death. The change of natriuretic peptide levels correlates with prognosis in acquired cardiovascular disease,^19, 20^ and the annual change of natriuretic peptide levels may also be associated with a poorer prognosis in ACHD.^10^ Our data substantiates this; ACHD patients prone to heart failure (such as patients with a systemic right ventricle or single ventricle) and patients with higher baseline BNP levels warrant serial BNP measurements and closer surveillance. If temporal BNP change is observed, earlier intervention (in the form of up titration of heart failure therapy, bringing forward haemodynamic intervention/s, and/or referral for advance heart failure assessment including transplantation) should be considered. The current study illustrates how an objective and simple marker may be well applied in the setting of a life-long chronic disease such as ACHD.

### BNP and its applicability to current ACHD and heart failure guidelines recommendations

It has been suggested that CHD may represent the original heart failure syndrome characterized by a triad comprising cardiac abnormality, exercise limitation, and neurohormonal activation,^11^ affecting younger patients compared to acquired heart disease cardiovascular cohorts. Different classes of biomarkers have been reported to be associated with adverse events in CHD, including markers of myocardial injury and/or inflammation. ^11, 21, 22^ Current guidelines for the management of ACHD state that serial testing of natriuretic peptides^14^ plays a role in identifying patients at risk for adverse events, and our current data unequivocally supports such an approach. Moreover, similar to acquired heart disease, we submit that ACHD patients on a heart failure trajectory should be monitored by serial natriuretic peptide levels according to the current American College of Cardiology (ACC)/American Heart Association (AHA) and European Society of Cardiology (ESC) guidelines for the management of heart failure. ^23 24^ It could also be of value to obtain a baseline natriuretic peptide level on all patients with ACHD, as even some patients with simple CHD lesions may encounter late complications,^25, 26^ so that comparisons can be made.

## Strength and limitations of the current study

To the best of our knowledge, this is the largest cohort study assessing BNP and its prognostic implication in ACHD. Our study is also the first study where temporal BNP change is shown to be prognostic beyond baseline BNP levels, independently of other established risk factors. Establishing the merits for serial BNP assessment in this population, we submit is a potential game-changer in ACHD clinical practice, namely to assess BNP as a reproducible, prognostic marker with ‘low cost’, easy to measure, and widespread availability.

Although our study provides solid representation of the ACHD spectrum, this remains a single-centre retrospective cohort study. We included patients who had BNP measurements only, as per study design and this might induce a selection bias towards patients with heart failure. Future, prospective studies of neurohormonal biomarkers may validate our data and show the impact of intervention on their levels and, thus outcome. Multicentre, prospective registries may shed additional light on the BNP as a surrogate marker in terms of diagnostics and prognosis of heart failure in ACHD and, in turn, whether BNP reduction improves prognosis in these patients.

## Conclusions

Baseline BNP measurements and temporal BNP change in our large, contemporary ACHD cohort were both significantly associated with all-cause mortality independently of underlying diagnosis, disease complexity, specific anatomic and haemodynamic features, including systolic ventricular function. The mortality of patients with BNP values in the highest quartile of measurements (>107 ng/L) was approximately 6-fold higher to the remaining patients. A similar effect was seen for change in BNP, where patients within the highest quartile of ΔBNP had 3.6-fold increased risk of death. BNP measurements are widely available, reproducible, simple and inexpensive to obtain, and should be incorporated in the routine, life-long surveillance of these patients.

## Data Availability

The data that support the findings of this study are available from the first author YY, upon reasonable request.

## Acknowledgements

None

## Figure Legends

**Figure 1. Boxplot of BNP and ΔBNP according to NYHA functional class and systemic ventricular function. A.** BNP and NYHA functional class. **B.** BNP and systemic ventricular dysfunction. **C.** ΔBNP and NYHA functional class. **D.** ΔBNP and systemic ventricular dysfunction.

**Figure 2. Scatterplot with regression line for the BNP or ΔBNP during follow up period. A.** Time point zero is denoted as the date of study inclusion and the first BNP measurement. **B.** Time point zero is denoted as the date when patients had primary endpoint or when they were censored without primary endpoint.

**Figure 3. Kaplan-Meier curves depicting survival free from the all-cause mortality according to the quartile of baseline BNP level and ΔBNP level. A.** Kaplan-Meier curves of survival free from all-cause mortality by the quartile of baseline BNP level. Shaded regions indicate 95% CI. **B.** Kaplan-Meier curves of survival free from all-cause mortality by the quartile of ΔBNP level. Shaded regions indicate 95% CI.

**Figure 4. Kaplan-Meier curves depicting survival free from the all-cause mortality according to baseline BNP level.** Kaplan-Meier curves of survival free from all-cause mortality by the higher BNP (>107 ng/L) and the lower BNP (≤107 ng/L). Shaded regions indicate 95% CI.

**Figure 5. Kaplan-Meier curves depicting survival free from the all-cause mortality according to ΔBNP level.** Kaplan-Meier curves of survival free from all-cause mortality by the higher ΔBNP (>35 ng/L) and the lower ΔBNP (≤35 ng/L). Shaded regions indicate 95% CI.

**Supplementary Figure 1.**

**A.** Distribution of BNP at baseline. **B.** Distribution of ΔBNP at serial assessment.

